# Unambiguous signatures of malignancies extracted from images of growing cells

**DOI:** 10.64898/2026.01.10.26343803

**Authors:** Gabriel Kalweit, Maria Kalweit, Wiktoria Checinska, Marijana Saric, Rebecca Berger, Elitsa Bodurova-Spassova, Justyna Rawluk, Nana Talvard-Balland, Anusha Klett, Marie Follo, Stefanie Kreutmair, Jesús Duque-Afonso, Michael Lübbert, Robert Zeiser, Joachim Frank, Roland Mertelsmann

## Abstract

As malignant transformation arises from dysregulated cellular programming with characteristic morphological changes, we hypothesized that cancer cells exhibit a unique morphological signature detectable in microscopy images of cells in vitro and in situ across modalities. To test this, we developed *CellSign*, an AI-based framework to generate Cell Dynamics Fingerprints, which (a) reconstruct morphological progression, (b) remove physiological variation, and (c) support automated malignancy assessment. To arrive there, cell morphology is represented by embeddings from vision foundation models, and a process we term Healthy-Component Reduction is used to refine these by subtracting normal physiological variation, thereby exposing residual disease-specific cues. Embeddings from healthy and malignant cells are organized with manifold learning and summarized with kernel density estimation. We show that unambiguous malignant signatures exist and that our method is robust across diverse datasets spanning breast cancer, lung cancer, and leukemia, transferring reliably between populations of single-cell images and multi-cell patches.

**Graphical Abstract:** 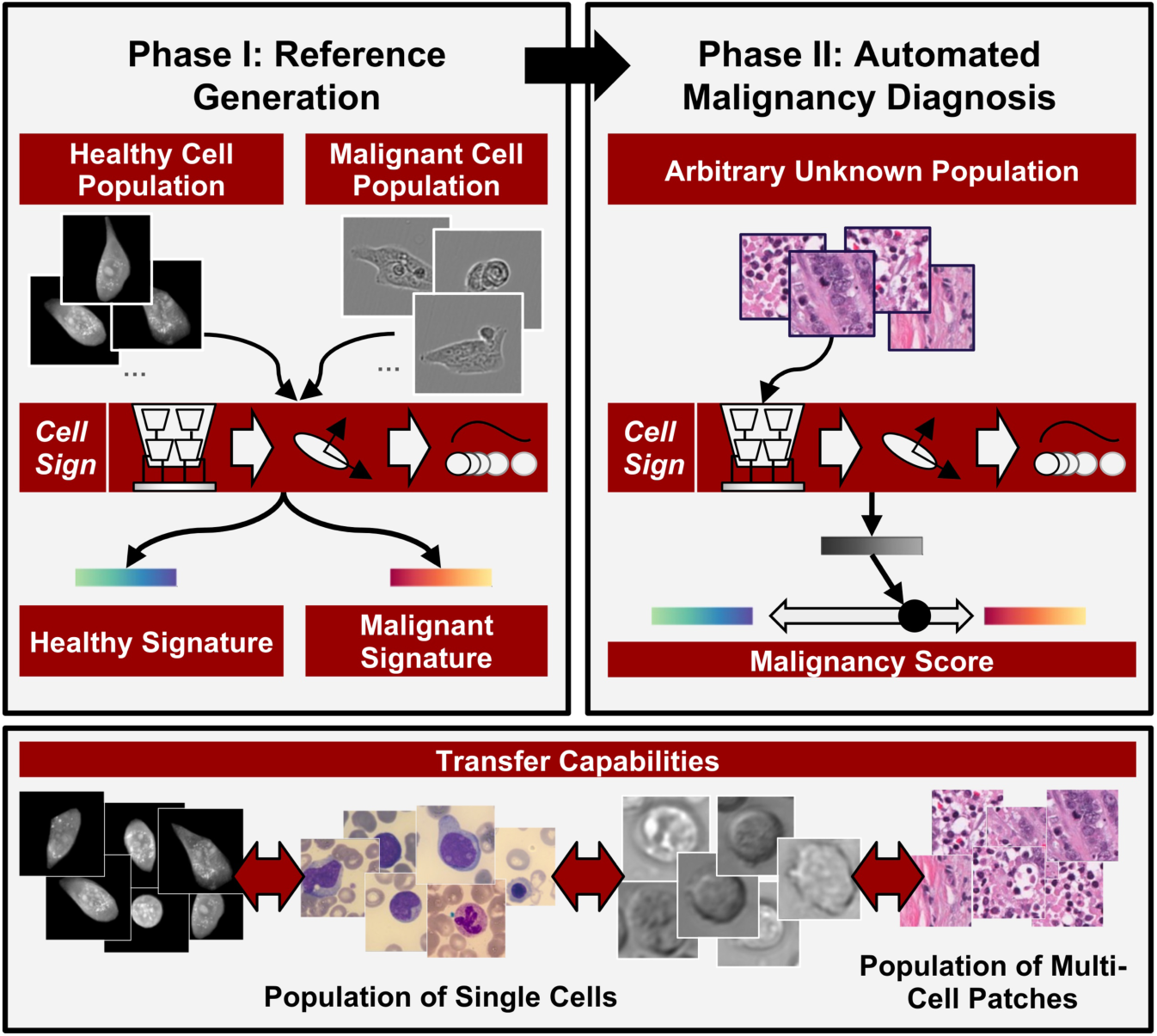

---

More than two decades after Douglas Hanahan and colleagues defined the hallmarks of cancer^1–3^, these principles continue to provide a unifying framework for understanding malignant transformation and cell fate. Cancer arises from the gradual acquisition of abnormal cellular behavior including hyper-proliferation, altered differentiation, and resistance to programmed cell death caused by mutations. Collectively, these mechanisms shape dynamic spatiotemporal cellular state transitions and drive tumorigenesis and malignant progression. While the underlying genetic and molecular principles of tumorigenesis are increasingly well characterized, their morphological manifestations on a cellular level remain elusive. Every process that regulates cellular plasticity and programming, such as (de-) differentiation, division, and programmed cell death is accompanied by structural change^4^. Some of these coordinated rearrangements leave visual morphological changes detectable by a microscope^38,40–42^. Even in healthy tissue, such programs generate rich morphological diversity. A cell’s size, nuclear texture, and cytoplasmic organization can reveal its position in the cell cycle or state of differentiation^5^. The same machinery that governs healthy cell function, cell cycle and maturation becomes persistently dysregulated during tumorigenesis. As a result, the structural logic of the cell changes (nuclei enlarge or deform, cytoplasmic organization becomes irregular, and population-level coherence) is lost^4,39^. We hypothesize here that malignant cell populations exhibit an unambiguous morphological malignancy signature that reflects cancer-associated biological processes. To uncover this signature, two complementary actions are required. One addresses the cancer signal within individual malignant cells. If we can identify and remove the morphological variation arising from non-malignant cell-cycle and maturation dynamics, the remaining structure should reveal a component that reflects morphological alterations linked to cancer, a *morphological malignancy residual*, in its pure form.

The second action addresses the structure inherent in the distribution of the whole cell population, since malignancy-related information can also emerge from the way a population of cells is distributed along the stages of the cell cycle and their maturation pathways. Recovering this progression requires organizing single-cell morphologies according to these underlying biological processes. Instead of relying on single-cell tracking over time, however, we hypothesize here that we can also reconstruct continuous morphological progressions from entire cell populations, where each snapshot of an unsynchronized sample reflects cells positioned along the cell cycle. The aforementioned malignancy residual along with the recovered population-level trajectory could yield representations of malignancy that are robust and generalizable across samples. To test this hypothesis, we take inspiration from structural biology, where unsupervised machine learning has been used to recover the conformational dynamics of biomolecules from ensembles of cryo-electron microscopy (cryo-EM) images^6,7^. In this setting, projections of molecules are captured in large collections of instantiations that differ subtly in the positions of mobile domains. By embedding these high-dimensional image representations into a manifold, researchers were able to order conformational states along continuous trajectories that describe molecular dynamics.

Following a similar rationale, we introduce *CellSign, a* framework to reconstruct morphological progressions from images of cell populations. This framework generates *Cell Dynamics Fingerprints (CDFs)*. Single-cell images are projected onto *cell embeddings* extracted by pretrained deep neural networks. Recent advances in deep learning^8–11^, particularly vision foundation models trained on large and diverse image corpora^12–19^, have made it possible to robustly detect and distinguish subtle morphological cell changes^20–25^. It has been shown^26^ on a per-cell basis that these embeddings are able to capture general morphological organization across contexts, learning transferable representations that may extend beyond specific stains or imaging modalities. Their capacity to encode both global and local structure could offer a robust and general foundation for distinguishing healthy physiological variation from disease-related change, and thus for isolating this morphological malignant residual.

We build upon the per-cell representations of Kalweit et al.^26^ to represent an entire population of cells. *Population* refers to a set of cells, originating from any of the following sources: blood smears, a specific solid tumor, a complete set of cells within a given dataset, or a sampled subset from that dataset. The overall procedure of the framework consists of two distinct phases. In the training phase, two reference CDFs are established, one for a healthy and one for a malignant population, each comprising asynchronously growing cells at different stages of maturation. In a subsequent diagnosis phase, when the reference CDFs are used for automated malignancy diagnosis of an unseen cell population, the CDF corresponding to the unseen population can be compared directly to the fixed references to obtain malignancy scores, without any need for retraining. We perform PCA on the cell embeddings of the healthy population to decompose the morphological structure and estimate the number of dominant healthy components, i.e., the number of components required to reach a defined explained variance threshold in the healthy subset. We project the malignant cell population embeddings onto the dominant healthy component subspace (*healthy residual*) and subtract this projection to obtain the residual morphology unexplained by normal variation, i.e, the *malignant residual*, in a process we call *Healthy-Component Reduction (HCR)*. After HCR on the malignant population, the healthy and the residual malignant ensemble instances are both organized in an unsupervised manner using the manifold learning approach PHATE^27^ (Potential of Heat-diffusion for Affinity-based Trajectory Embedding) to reconstruct continuous morphological trajectories and extract a *pseudotime* ordering that reflects progressive morphological change attributable to the cell-cycle and maturation progression. The population-level distributions are then summarized through kernel density estimation (KDE) to obtain reference CDFs for the healthy and the malignant population.

Once the training stage of the automated malignancy diagnosis is complete, the learned references can serve as a fixed basis for diagnosing new, previously unseen populations without any additional training, hence they are archived for general use. Populations to be diagnosed are embedded, sorted, and pooled into CDFs in the same manner. Their similarity (in terms of an appropriate metric) to the healthy or malignant reference CDFs defines a malignancy score, which places each sample population along a continuous morphological axis between healthy and malignancy states. An overview of *CellSign* is given in Fig. 1. We demonstrate the validity and generality of this approach across sixteen distinct datasets, spanning diverse imaging modalities, biological sources, and levels of supervision. These include solid cancer datasets, such as solid epithelial cell lines MCF10A and JIMT-1, and histological slides from breast and lung tissue. Further, they contain hematological live-cell recordings obtained with brightfield and LED-array microscopy, stained white blood cell samples, and primary acute myeloid leukemia (AML) patient specimens. Across these varied contexts, we found that CDFs with HCR (HCR-CDFs) consistently capture a shared morphological malignant signature that is unambiguous and stable across disease types and patients, and transferable among imaging platforms as well as between sets of single-cell images and sets of multi-cell patches.

**Fig. 1.**
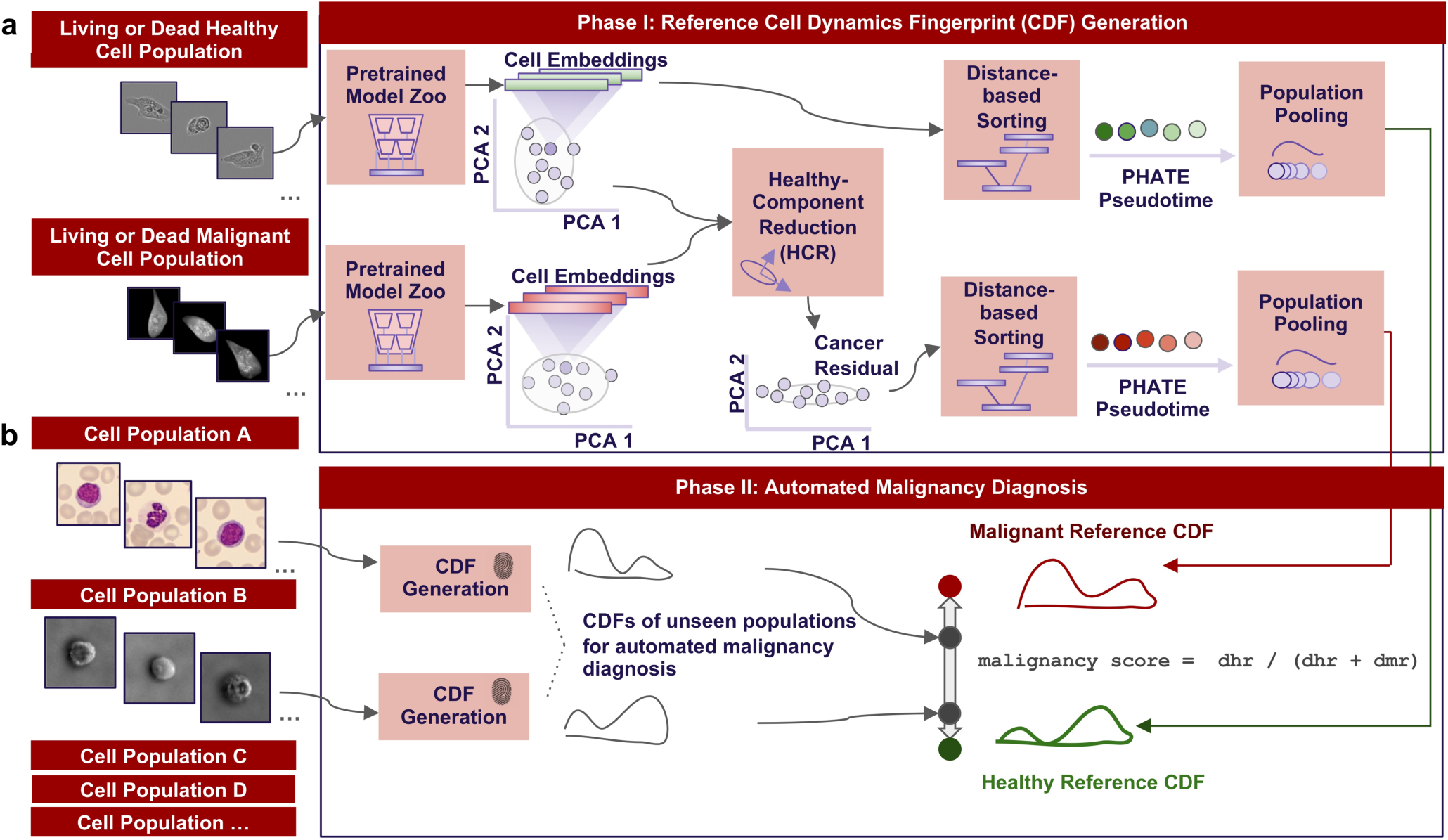
Overview of Cell Dynamics Fingerprints (CDF) with Healthy-Component Reduction (HCR). **a**, Foundation model embeddings are generated from healthy and malignant single-cell image datasets using a pretrained deep vision model zoo. In the training phase, we perform HCR, where principal component analysis (PCA) is applied to the embeddings of the healthy population to identify dominant morphological variance components as healthy residual, which is then removed from the cell embeddings of the malignant population to obtain the malignant residual. Both healthy embeddings and the malignant residual are ordered along a low-dimensional manifold using PHATE^27^, yielding a pseudotime ordering that reflects continuous morphological progression. Kernel density estimation (KDE) over this pseudotime ordering produces smooth reference CDFs for a healthy and a malignant population. **b**, To perform automated malignancy diagnosis in the second phase, cell populations are processed through the same PHATE-KDE procedure. New, unseen CDFs are then compared with the reference CDFs to quantify a malignancy score, where *dhr* and *dmr* denote distances (here: using the Wasserstein distance^43^) to the healthy and malignant references, respectively.

## Results

### HCR Removes Physiological Morphological Variation, and PHATE Reveals Continuous Dynamics in JIMT-1 Cells

We examined the related breast epithelial cell lines JIMT-1 and MCF10-A (Fig. 2a) to assess the behavior of the malignant residual under controlled biological conditions. Brightfield recordings of the breast cancer line JIMT-1, treated with Raptinal and observed over time until apoptosis, served as the malignant population, while the non-malignant epithelial line MCF10-A provided the healthy reference. For both populations, single-cell images were extracted at different times using deep segmentation with CellSAM^28^. Each single-cell image was represented by concatenated embeddings^26^ derived from feeding the image into multiple vision foundation models (we used a combination of DINOv2^13^, ConvNeXt^14^ and Swin^15^), forming the cell embeddings. The principal component and subspace-angle analyses (Fig. 2b) show the expressiveness of the cell embeddings and the degree to which HCR isolates disease-related variance from shared morphological structure. The stacked eigenvalue spectra (Fig. 2b, left) reveal for the healthy, cancer, and residual embeddings a structured space with pronounced peaks in the leading components. The cumulative explained variance and participation ratio of 17.2 for the cancer embeddings (Fig. 2b, middle) indicate that most of the variance is concentrated in a few dominant components before HCR. By removing these shared components, HCR substantially reduces variance in the leading axes. Principal angle analysis (Fig. 2b, right) confirms this effect: while the cancer and residual subspaces still show partial overlap (mean angle ≈35°), the residual becomes nearly orthogonal to the healthy subspace (mean angle ≈90°), indicating that HCR effectively removes healthy-dominated morphological variance.

**Fig. 2.**
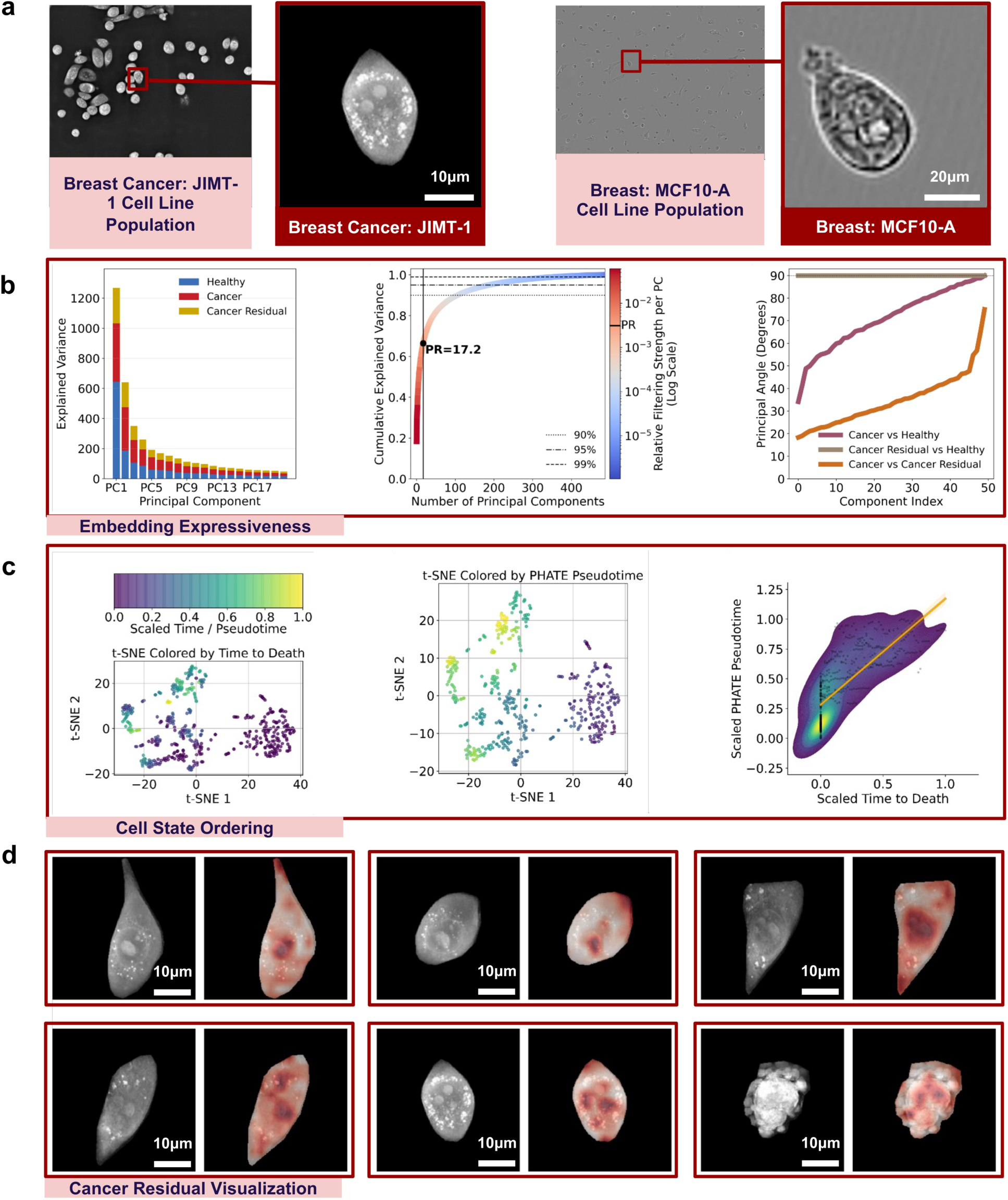
PHATE ordering and HCR reveal continuous malignancy-associated morphological dynamics. **a**, Representative images of breast epithelial cell line populations (left) and extracted single cells (right) used as healthy (MCF10-A, 79,178 cells) and malignant (JIMT-1, 487 cells) references. **b**, Principal component analysis of concatenated foundation model embeddings (DINOv2, ConvNeXt, Swin) after HCR. The stacked eigenvalue spectra (left) show reduced contribution from the leading principal components, as reflected in the cumulative explained variance plot with participation ratio (middle; color indicates reduced variance after HCR). Principal angle analysis (right) compares subspaces of healthy, cancer, and residual embeddings, with mean angles of 35° between cancer and residual and 90° between healthy and residual. **c**, PHATE manifold embedding of single-cell embeddings reconstructs a continuous trajectory from the population for JIMT-1 cells. t-SNE projections are colored by time to death (TTD, left) and PHATE-derived pseudotime (middle).

To test whether the foundation model embeddings capture biologically meaningful progression within the malignant population, we next applied PHATE to JIMT-1 embeddings (Fig. 2c). The resulting pseudotime ordering reconstructed a continuous trajectory along a morphological manifold that closely aligned with the time to death (TTD) of the cells (Pearson r = 0.78, Spearman ρ = 0.85). The TTD was determined by tracking single cells over time and measuring the interval until the first labeled apoptotic signs (such as membrane blebbing). This correlation indicates that morphological variation within the manifold encodes a coherent temporal sequence of cell-state changes. Finally, residual heatmaps (Fig. 2d) visualize the spatial localization of deviations from the healthy morphological subspace. Higher-residual magnitudes, shown in red, correspond to regions exhibiting cancer-associated structural alterations, which are consistently concentrated in nuclear and perinuclear compartments.

### HCR-CDFs Enable Robust Transfer from Live-Cell Microscopy to Tissue Sections Across Cancer Types

We next evaluated how well the HCR-CDFs perform when transferred across different imaging modalities and tissue types to test the generalizability of their signatures (Fig. 3a). First, we created reference HCR-CDFs on the breast epithelial cell lines JIMT-1 and MCF10-A. For automated malignancy diagnosis, we used unseen CDFs from tissue sections of breast (BCSS-WSSS) and lung cancer (LUAD). These domains form a controlled progression from less to more complex due to the shift in tissue type, allowing us to test whether the disease-specific morphological signature generalizes across scales and modalities.

**Fig. 3.**
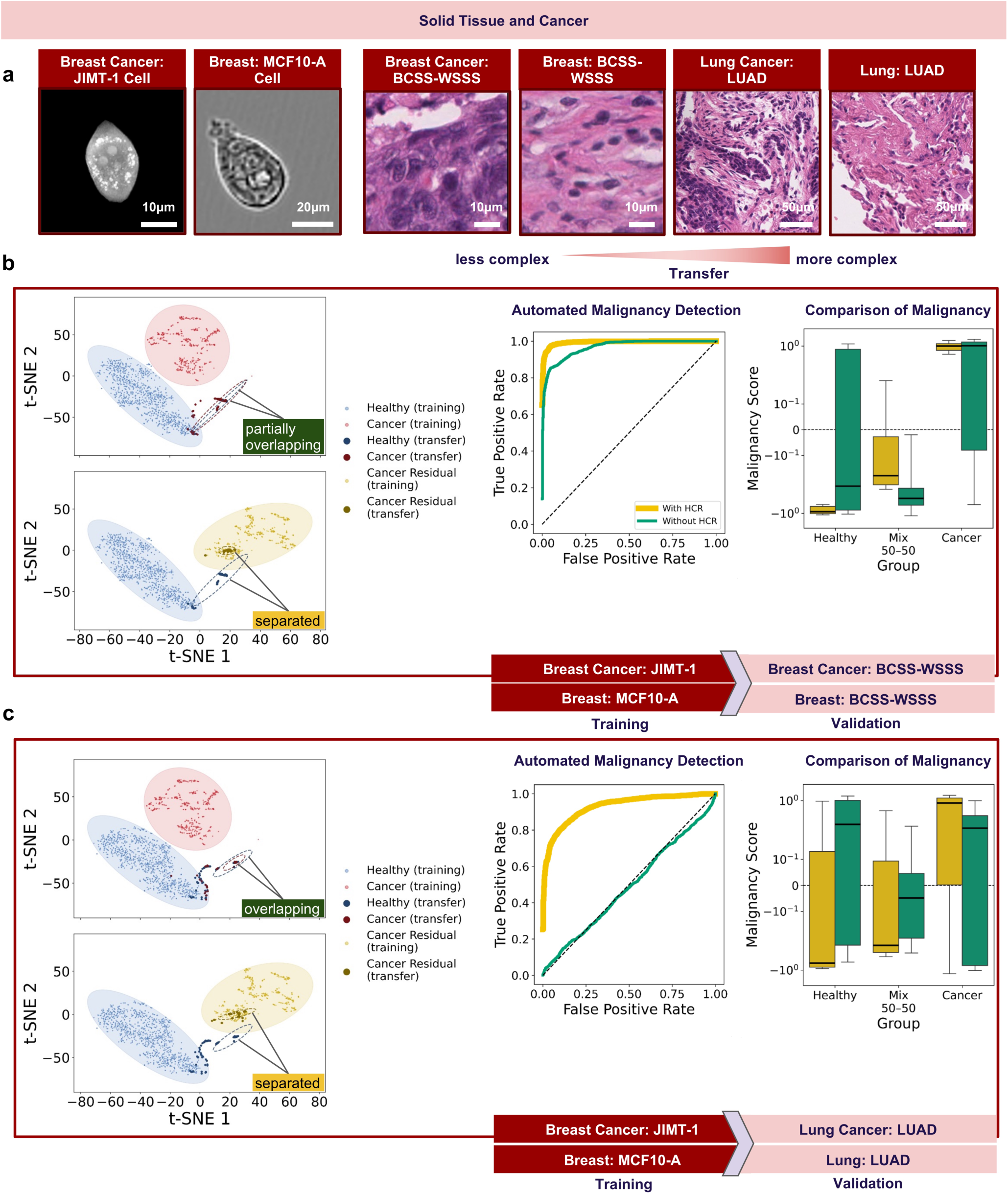
HCR enhances cross-modality and cross-tissue generalization. **a**, Representative image pairs showing increasing visual complexity from single brightfield cells (JIMT-1, 487 cells; MCF10A, 79,178 cells) to solid tissue sections (breast BCSS-WSSS, 23,422 patches; lung LUAD, 16,678 patches). **b-c**, Left: Two-dimensional representation of the training embeddings (transparent) and the projection of the transfer embeddings (bold). Ellipses (colored by group) outline the 95% confidence contour of a Gaussian fitted to each group’s embeddings (solid: training, dashed: transfer). Middle: Median ROC curves of automated malignancy detection across 20 runs with and without HCR for CDFs trained on JIMT-1 (malignant) and MCF10A (healthy) brightfield images and evaluated across domains. Right: Boxplots showing the distribution of estimated malignancy scores for homogeneous healthy, mixed, and malignant populations across 20 runs. **b**, Cross-modality transfer from brightfield to breast histopathology (BCSS-WSSS) shows consistent performance (macro median AUROC = 0.994 with HCR vs. 0.968 without HCR). **c**, Cross-tissue transfer from breast to lung histopathology (LUAD) shows improved performance with HCR (macro median AUROC = 0.927 with HCR vs. 0.504 without HCR).

Correlation analysis (right) between pseudotime and TTD (r = 0.78, ρ = 0.85) shows their correspondence. **d**, Residual heatmaps for JIMT-1 cells show the norm of the residual signal after HCR (right image of each pair) next to the original brightfield cell image (left). Higher residual magnitudes (dark red) mark structural deviations from the healthy morphological subspace, seen mainly in nuclear and perinuclear regions.

Two-dimensional projections using t-distributed Stochastic Neighbor Embedding^29^ (t-SNE; Fig. 3b and 3c, left) allow visual inspection of the embeddings before and after applying HCR. For each run, we computed HCR-CDFs by sampling 1,000 healthy cells and 1,000 malignant cells. From these, we drew 30 random subpopulations of 200 cells each with replacement, computed a fingerprint for each subpopulation, and averaged them to obtain the mean healthy and mean malignant reference fingerprints. For evaluation, we analogously sampled 1,000 populations of 200 cells from the malignant and healthy transfer test sets, both homogeneously and at a pre-set 50-50 ratio. We then derived a malignancy score that quantifies how closely a population’s fingerprint aligns with the malignant or healthy reference fingerprints. The score is based on the similarity (here in terms of the Wasserstein distance) of each population’s fingerprint to the healthy and malignant reference CDFs, where higher values indicate stronger similarity to the malignant reference. Automated malignancy diagnosis was evaluated using the macro-median receiver operating characteristic (ROC) curves and their corresponding macro-median area under the curve (AUROC) values derived from these malignancy scores, with reported AUROCs representing the median performance across 20 runs.

When transferring from the cell lines to breast histopathology from the BCSS-WSSS dataset (Fig. 3b), the cell embeddings show ambiguity, as healthy and malignant cells partially overlap before application of HCR. This is evident from the overlapping dashed ellipses, which represent the 95% confidence contours of Gaussian fits to the healthy and malignant transfer embeddings. We hypothesize that the partial overlap arises from a distribution shift when moving from live-cell brightfield to stained tissue imaging, as well as from the difference in spatial scale between single-cell images and multi-cell tissue patches. The high degree of going out of distribution leads to histopathology patches clustering tightly in the feature space, because the predominant non-malignant specific cues extracted from label-free images (training) do not remain sufficiently expressive when applied at patch scale and under staining conditions (transfer). Together, these factors weaken consistent class separation after transfer. Applying HCR removes this ambiguity by filtering out modality-induced variation, restoring alignment with the training-domain structure. After HCR, the malignant residuals become clearly separated from the healthy population, indicating that the disease-specific structure is successfully recovered. As a result, performance remains high with HCR (macro AUROC = 0.994) and remains good without HCR (0.968), showing that cell dynamics alone already supports discrimination of malignancy, while HCR further improves separation and avoids domain-shift-related effects.

A more challenging transfer to lung adenocarcinoma tissue (LUAD) is shown in Fig. 3c, where the dashed ellipses reveal an even higher degree of overlap. The red dashed ellipse representing malignant embeddings is fully contained within the blue healthy dashed ellipse, indicating complete loss of separability before applying HCR. This substantial ambiguity results from the combined effects of changed tissue architecture and the smaller apparent magnification of cells in LUAD tissue sections compared to the breast dataset, introducing an additional multi-scale shift, causing transfer embeddings to cluster even more tightly. Applying HCR resolves this ambiguity and produces clear separation: malignant residuals align precisely with the training-domain disease structure, while healthy embeddings remain distinct. Consistently, quantitative performance improves markedly with HCR (AUROC = 0.927 vs. 0.504 without), confirming that application of HCR uncovers disease-specific information even across major modality, scale, and tissue shifts.

### HCR-CDFs Enable Cross-Cell-Line Transfer in Live-Cell Brightfield and Robust Generalization to Stained Microscopy in Blood

We next examined whether the approach extends to hematological malignancies (Fig. 4a). Using the Kreutmair AML dataset comprising seven leukemia cell lines (KASUMI-1, MKPL-1, MOLM-13, OCI-AML2, OCI-AML3, OCI-M2, THP-1) and healthy PBMCs, we evaluated CDFs with and without HCR under complementary settings. In the embedding space (Fig. 4b, left), both training and validation data show substantial ambiguity, as healthy and leukemic populations partially overlap. Since all images were acquired under identical conditions and with the same microscope, the overlap reflects biological similarity rather than clustering due to imaging modality. Applying HCR isolates the malignant residual, which separates clearly from the residual of the healthy population, demonstrating that HCR resolves intrinsic morphological ambiguity and disentangles disease-related similarity structure within a single imaging domain.

**Fig. 4.**
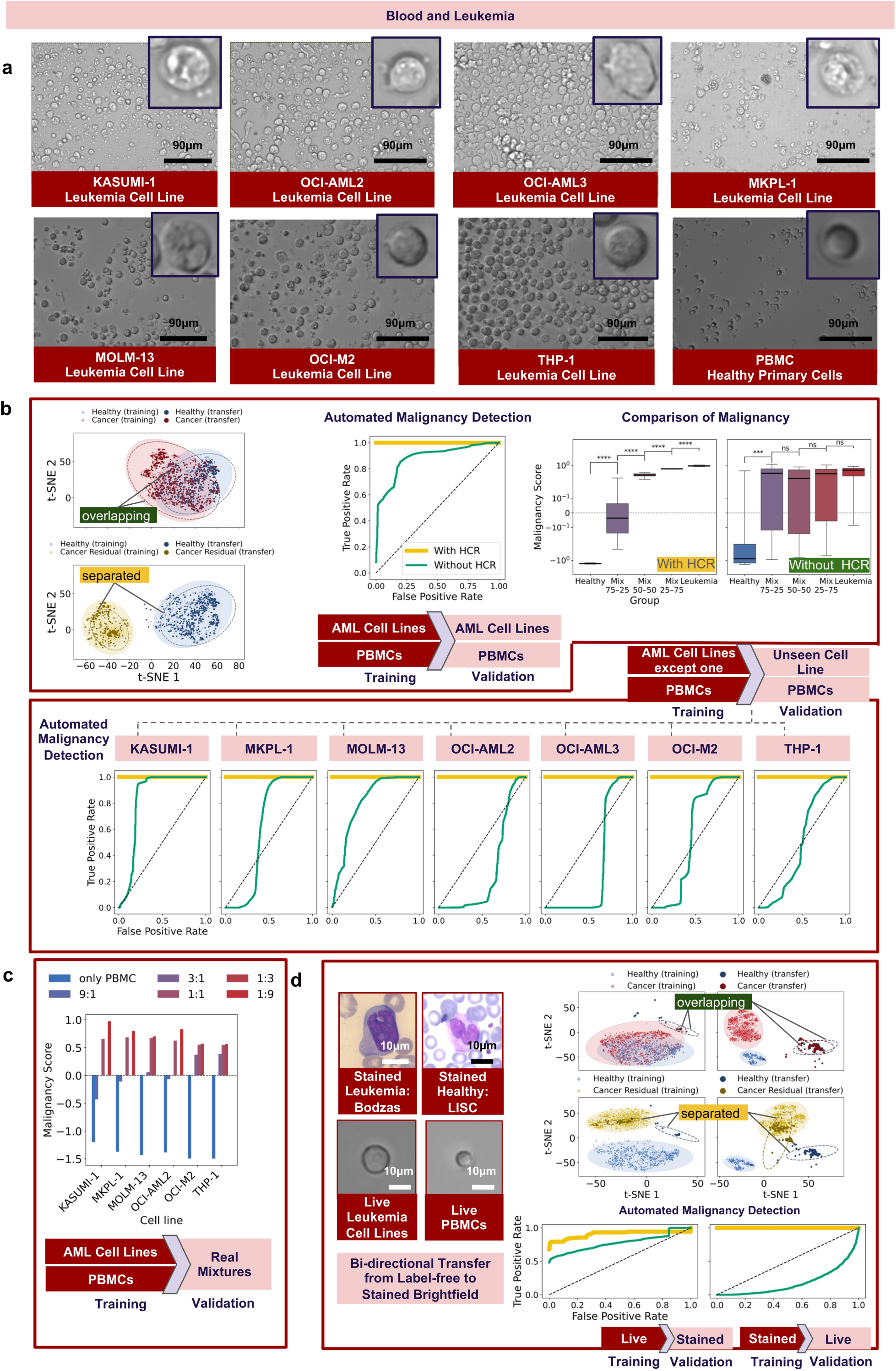
HCR-CDFs enable cross-cell-line and cross-modality generalization in live-cell and stained microscopy in blood. **a,** Representative original brightfield images of leukemia cell lines (KASUMI-1, MKPL-1, MOLM-13, OCI-AML2, OCI-AML3, OCI-M2, THP-1; ∼9,000-20,000 cells per cell line) and healthy PBMCs (24,000 cells). Insets show examples of automatically extracted single-cell crops. **b**, Top left: t-SNE visualization of training and transfer embeddings. Top middle: Performance of automated malignancy detection using reference CDFs containing all AML lines and PBMCs with (yellow) and without (green) HCR on a separate validation set. Top right: Malignancy score distributions across mixed populations. Bottom: ROC curves of a leave-one-cell-line-out transfer, where malignant reference CDFs are built from all but one AML cell line. **c**, Malignancy scores for each AML line and for real PBMC-AML co-culture mixtures at different PBMC:AML ratios, along with the healthy PBMC reference. **d**, Cross-modality transfer between live-cell brightfield and stained blood smear images. t-SNE visualizations show training and transfer embeddings for both transfer directions (stained to live and live to stained; stained samples are from Bodzas, 5,091 malignant cells, and from LISC, 257 healthy cells). Reference HCR-CDFs for label-free AML and PBMC generalize to stained leukemic and healthy white blood cells (macro AUROC ≈ 0.91 with HCR vs. 0.76 without), and vice versa (macro AUROC ≈ 1.00 with HCR vs. 0.18 without).

To calculate the performance of the automated malignancy detection, for each run, we computed HCR-CDFs as described above by sampling 1000 cells per population (healthy and malignant) and generating 30 subpopulations of 200 cells each. Evaluating on a separate validation set (within the training domain), CDFs derived from all AML lines and PBMCs achieved near-perfect discrimination between healthy and leukemic populations (macro AUROC ≈ 1.00 with HCR, Fig. 4b, top middle, ROC-AUC curves), confirming that the embeddings capture consistent disease-related morphology. Synthetic mixtures (75–25%, 50–50%, 25–75%) created by combining healthy and leukemic cell embeddings in defined proportions revealed a smooth progression of malignancy scores (Fig. 4b, top right, boxplot with HCR) from healthy to mixed to leukemic groups, preserving clear group separations (**** p < 0.0001). Without HCR, overall discrimination decreased (macro AUROC = 0.870, Fig. 4b, top right, boxplot without HCR), and the consistency of the malignancy scores dropped substantially, leaving no significant group separations, a result which is consistent with the visual ambiguities observed in the embedding space. To test whether the performance keeps stable when we generalize to CDFs of unseen AML cell lines, we performed a leave-one-cell-line-out transfer, where the malignant reference CDFs was computed on all cell lines but the one that was validated on (Fig. 4b, bottom). In this setting, HCR consistently improved cross-cell-line discrimination, yielding stable malignancy detection performance across all subtypes (average median AUROC = 1.00 with HCR vs. 0.56 without).

We next examined whether these patterns could be confirmed using real co-culture mixtures in which AML and PBMC cells were plated together in defined ratios in the same well (Fig. 4c). Malignancy scores were computed for each co-culture mixtures at the defined ratios PBMC:AML of 9:1, 3:1, 1:1, 1:3 and 1:9. For healthy CDF reference we used the already computed PBMC reference, since the real co-culture mixture recordings did not contain purely healthy samples. Across all AML lines, the malignancy scores increased proportionally with the leukemic fraction, confirming that the CDFs capture differences in cellular composition quantitatively.

Finally, we evaluated whether HCR-CDFs enable robust transfer to stained microscopy in blood (Fig. 4d). Visual inspection of the two-dimensional projections showed that both training and transfer embeddings initially exhibited substantial overlap between healthy and leukemic populations, indicating that modality-specific variation blurs disease-related structure. Applying HCR isolated the malignant residual relative to the healthy reference, yielding distinct, well-separated clusters across modalities.

Performing automated malignancy detection, reference HCR-CDFs from healthy and malignant label-free live-cell recordings of the Kreutmair dataset successfully generalized to CDFs of stained white blood cell (Bodzas and LISC) images (macro AUROC ≈ 0.91 with HCR vs. 0.76 without). The reverse transfer from Bodzas and LISC to Kreutmair showed an even stronger difference (macro AUROC ≈ 1.00 with HCR vs. 0.18 without). Without HCR, discrimination decreased, especially in the Bodzas+LISC-to-Kreutmair transfer, indicating that modality-specific variance impairs generalization when the healthy component is retained. In contrast, application of HCR maintained high separability in both directions, confirming that disease-specific morphological structure is preserved across modalities.

### HCR-CDFs Enable Robust Transfer even in Case of Small Reference Data and Extreme Distribution Shift

To evaluate the robustness of HCR-CDFs in case of severe distribution shifts, we assessed their transferability across distinct biological and imaging domains (Fig. 5a). In the first experiment (Fig. 5b, left), CDFs were extracted from all (pathological and non-pathological) cells of a single AML patient from AML Dataset 2 together with stained healthy white blood cells from the Raabin dataset. Despite the malignant reference being derived from a single-patient blood smear, CDFs retained high discriminative power across an independent AML Dataset 1 with a patient cohort of 25 patients and healthy Acevedo samples when using HCR (macro median AUROC = 0.93 with HCR vs. 0.74 without). This result demonstrates that HCR restores generalizable disease-specific information even when the malignant reference is highly limited in diversity (in this case consisting of only one patient sample).

**Fig. 5.**
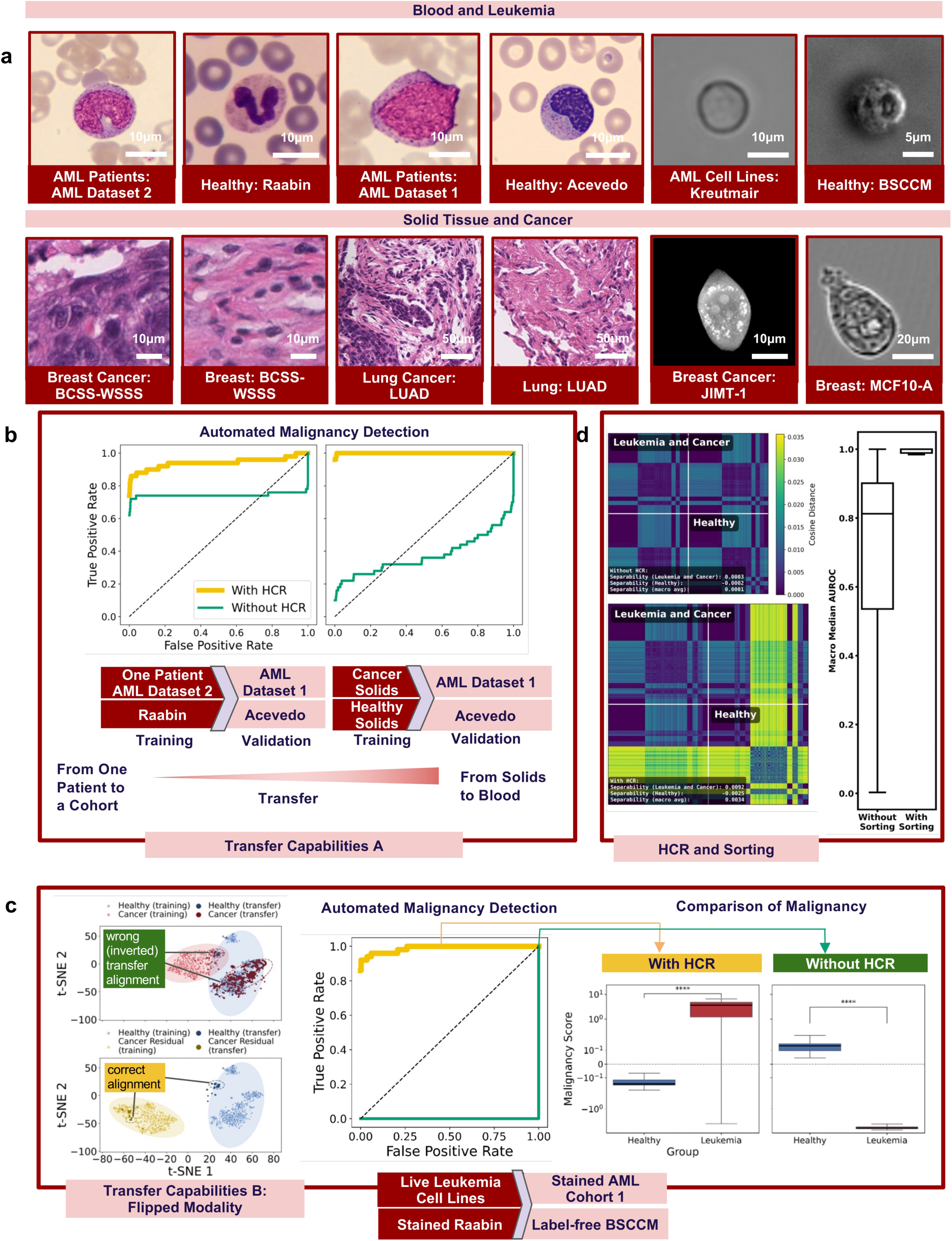
HCR enables robust transfer in case of small reference data and distribution shift. **a,** Representative examples of datasets spanning hematologic and solid tumors across stained and label-free imaging modalities. Top: Brightfield images of AML patient samples (AML Dataset 1 and 2, ∼200-400 cells per patient), healthy white blood cells (Raabin, 16,633 cells; Acevedo, 17,092 cells), and label-free LED-array recordings (BSCCM, 9,453 cells). Bottom: Stained histopathology of breast (BCSS-WSSS, 23,422 patches) and lung (LUAD, 16,678 patches) cancers, and brightfield or live-cell images of JIMT-1 (487 cells) and MCF10-A (79,178 cells). **b**, ROC-AUC curves for transfer across domains. Left: Transfer from a single AML patient (∼200-400 cells) from AML Dataset 2 to the AML Dataset 1 and healthy Acevedo cells. Right: Transfer from solid-tumor datasets (breast and lung) to blood-based cohorts. **c**, Flipped-modality transfer (with HRC: AUROC = 0.98 and without HCR: AUROC = 0.0), in which both staining and imaging modality are inverted between training and validation (stained vs. label-free). Right: Malignancy score distributions for the corresponding comparisons across modalities. **d**, Wasserstein-distance matrices between leukemia/cancer and healthy fingerprints (left) and comparison of macro median AUROCs of all experiments with and without sorting via PHATE (right).

In the second experiment (Fig. 5b, right), automated malignancy detection using reference CDFs built from all solid-tumor datasets, including breast (JIMT-1, MCF10-A, BCSS-WSSS) and lung (LUAD) data, was evaluated on hematological patient samples. Without HCR, transfer performance was poor (macro AUROC = 0.37), reflecting strong modality and tissue bias. Applying HCR re-established separability between healthy and malignant fingerprints, raising the macro AUROC to 1.00 and confirming that disease-specific morphological variance can transfer robustly across biological systems. The most stringent test involved a flipped-modality configuration (Fig. 5c), in which both staining and imaging domains were inverted between training and validation. CDFs were built on label-free AML cell lines and stained healthy cells, then evaluated on stained AML samples from AML Dataset 1 and label-free healthy cells. Without HCR, this inversion led to complete failure (macro AUROC = 0.0), indicating an inverted classification dominated by irrelevant visual cues such as color or illumination. Applying HCR restored biologically meaningful alignment and achieved near-perfect discrimination (macro AUROC = 0.98), demonstrating that the residual representation remains interpretable even when imaging contrast is reversed. The boxplots illustrate that HCR preserves a clear separation between healthy and leukemic groups, whereas without HCR, the alignment polarity is reversed, indicating that the embeddings no longer reflect biologically meaningful structure. The corresponding two-dimensional projections confirm that HCR consistently isolates the malignant-associated residual across modalities.

Across all experiments and datasets, Wasserstein-distance matrices between cancer and leukemia versus healthy CDFs (Fig. 5d) showed that HCR restores distinct block structures separating malignant from healthy states across tissues and imaging modalities (34-fold increase in separability compared to without HCR). Moreover, comparison of macro median AUROCs across all experiments with and without PHATE-based sorting revealed a consistent improvement when temporal ordering was included, indicating that pseudotime organization enhances both performance and interpretability of morphological CDFs.

## Discussion

Our results indicate that unambiguous morphological signatures of malignancy exist and that we found a robust way to identify them across datasets. We did so by introducing a unified framework, *CellSign*, for capturing disease-associated morphology directly from cell population images across scales, modalities, and biological systems. It combines foundation model embeddings to represent single-cell images with HCR to separate disease-related variance from physiological background variations, orders the resulting embeddings with PHATE to capture morphological progressions, and pools them using KDEs to form CDFs. Across all tested settings, from breast epithelial lines to hematological malignancies, HCR-CDFs consistently preserved discriminative morphology while suppressing modality- and dataset-specific bias. Here, we used *CellSign* to separate cancerous from non-cancerous signal; however, we strongly believe that the utility of this framework also extends to other entities.

PHATE ordering recovered continuous morphological trajectories from populations of asynchronously imaged JIMT-1 breast cancer cells, where each snapshot reflects a different position along the continuum of the cell cycle and differentiation. This continuity aligns with experimentally measured temporal progressions, indicating that the embeddings encode underlying physiological dynamics such as cell-cycle related variation. HCR showed to effectively remove these recurring physiological morphological patterns, isolating a residual subspace that captures disease-specific structure. Beyond quantitative separability, the approach also enables visual explanations for which morphological features may drive these differences, highlighting where and how morphology departs from normal physiology.

Although these early visualizations represent only an initial step toward morphological interpretability, they open the possibility of systematically tracing the visual hallmarks of malignancy recognized by pathologists in an objective, reproducible, data-driven manner.

A key advance of this study is the generalization of morphological fingerprints across spatial and biological scales. The same embedding and reduction strategy applies to populations of single-cell images and even to multi-cell histopathology patches. This bidirectional scalability, between the single-cell and tissue levels, enables seamless translation of information between experimental systems and clinical samples. The ability to transfer from cell lines to patient histology, and even from brightfield to stained microscopy, demonstrates that HCR-CDFs capture morphology in a representation that is both context- and modality-agnostic. These results outline a path toward more standardized and accessible morphology-based diagnostics. Because morphological fingerprints can be computed directly from conventional microscopic images, they could, in the future, support analyses of routinely collected patient samples, such as blood smears. Detecting subtle shifts in population-level fingerprints may eventually contribute to earlier recognition of malignant transformations, complementing existing diagnostic procedures. The impact of individual oncogenes introduced into non-malignant cells on the recognition as a cancer cell by *CellSign* is currently investigated. Future work should clarify the biological origins of the residual morphological signal and determine which aspects of cellular organization contribute most strongly to the disease-specific fingerprint. Linking the learned residuals to molecular markers, spatial transcriptomic data, or mechanical properties could reveal the underlying pathways that connect visual morphology to functional dysregulation.

## Data Availability

All data produced in the present study are available upon reasonable request to the authors.

## Authors Contributions

The project was conceived and the experiments were designed by GK, MK, JF, and RM. GK and MK implemented the method and performed the computational analyses. EBS, AK, and MF generated, recorded, and annotated the apoptosis dataset for this study. MS, WC, and SK designed, generated and recorded the leukemic cell line datasets for the study. RB, JR, and JDA generated and recorded the stained AML patient datasets for use in the study. NTB and RZ designed and generated datasets supporting ongoing investigations into the impact of individual oncogenes introduced into non-malignant cells. The manuscript was written by GK, MK, JF, and RM, with conceptual input from ML and RZ. All authors reviewed the manuscript.

## Acknowledgements

This research was funded by the Mertelsmann Foundation by the grant numbers Nr. 2025031 (to. JD-A) and MF-2024017 (to SK). SK was further supported by a fellowship for attending physicians from the Promedica Foundation.

## Competing Interests

RM, GK, MK, and JF are inventors on a patent application related to the method described in this manuscript (Patent application DE 10 2025 150 856.9; applicant: BioThera Institut GmbH; status: filed). The patent covers the computational procedure for extracting and comparing morphological signatures from microscopy images. RZ received speaker fees from Novartis, Therakos, Sanofi, medac, Incyte and Neovii. J.D-A. received speakers honoraria from Roche, Amgen, Riemser, SOBI, IPSEN, Abbvie, Beigene, NovoNordisk, AstraZeneca and Lilly and travel support from Abbvie, Lilly, Roche, Gilead, IPSEN, SOBI, and Beigene. All other authors declare no competing interests.

## Methods

### Datasets

We evaluated *CellSign* across a comprehensive range of imaging modalities, tissues, and disease contexts (Fig. 6).

**Fig. 6.**
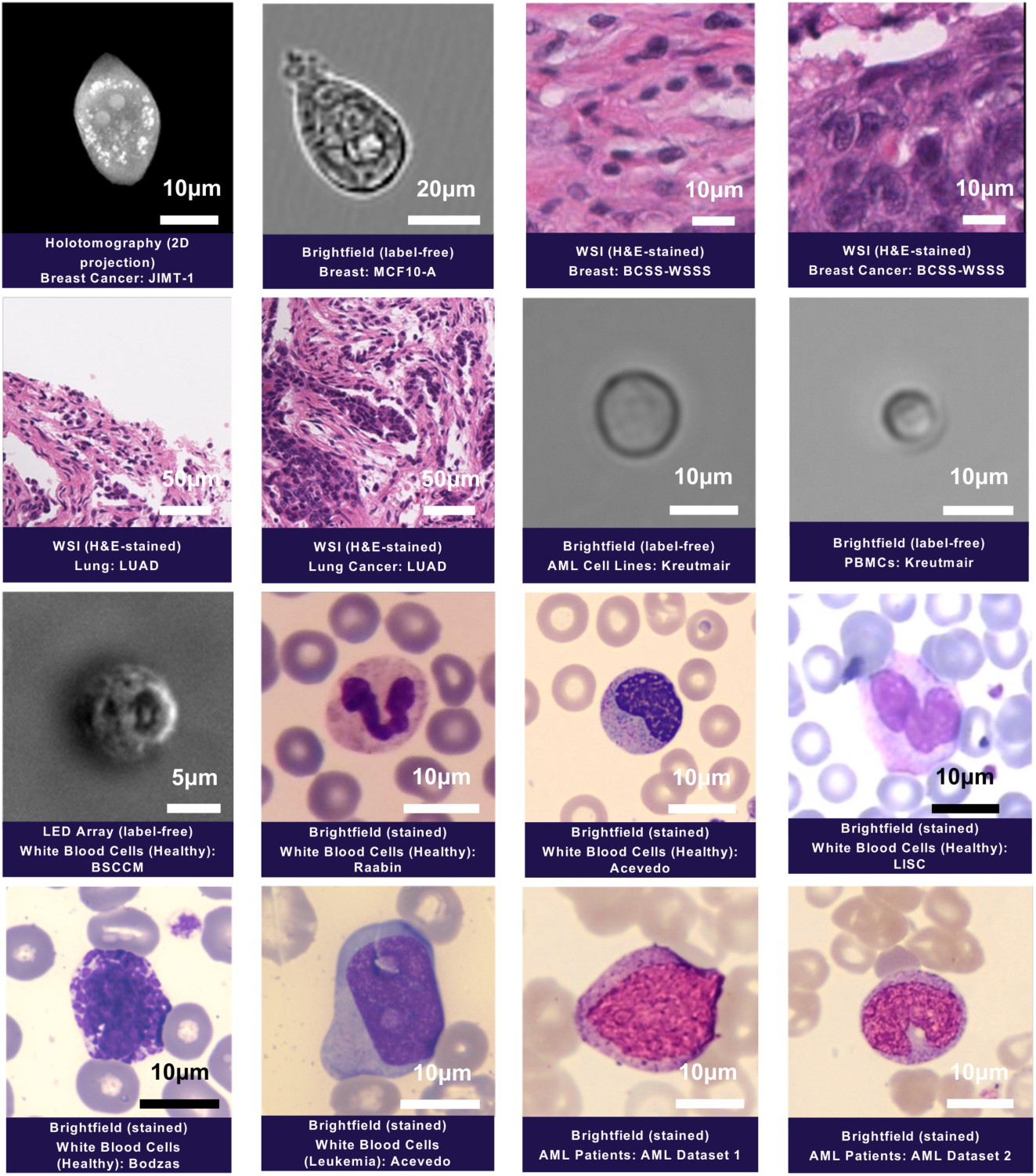
Datasets used in this study. Representative examples of imaging modalities and specimen types included in this work. Top row: label-free holotomographic projections of breast cancer cells (JIMT-1) and brightfield images of non-cancerous breast epithelial cells (MCF10A), followed by hematoxylin-eosin (H&E)-stained whole-slide images (WSIs) of breast cancer (BCSS-WSSS). Middle row: H&E-stained WSIs of lung cancer (LUAD) and label-free brightfield images from acute myeloid leukemia (AML) cell lines and healthy peripheral blood mononuclear cells (PBMCs). Bottom rows: LED-array label-free and brightfield-stained images of healthy white blood cells from multiple sources (BSSCM, Raabin, Acevedo, LISC) and stained leukemic and AML patient samples from Bodzas, AML Dataset 1, and AML Dataset 2.

#### Own datasets

Label-free live-cell datasets included holotomographic 2D projections of breast cancer cells (JIMT-1, n = 487). The JIMT-1 images^26^ correspond to 15 individual cell tracks that were annotated by an expert until apoptosis. The images are 2D projections of 3D volumes. JIMT-1 cells were cultivated using Dulbecco’s modified eagle medium (DMEM) FluoroBrite (Gibco), supplemented with 10% fetal bovine serum (FBS; Gibco), 1x L-glutamine (Gibco), and 1% Pen Strep (10,000 Units/ml penicillin, 10,000 µg/ml streptomycin; Gibco). JIMT-1 cells were treated with 2.5 µM, 5 µM and 10 µM of Raptinal (Sigma Aldrich) for 24h to induce cell death. 300 nM of Biotracker Apo15 (Sigma) was used as a fluorescence marker to detect early stage apoptosis. The cells were seeded in a µ dish 35 mm, low glass bottom (Ibidi). Brightfield Images were acquired every 15 min and fluorescence every 3h.

Label-free brightfield images from the Kreutmair dataset included seven AML cell lines (KASUMI-1, MKPL-1, MOLM-13, OCI-AML2, OCI-AML3, OCI-M2, THP-1) and healthy PBMCs, comprising ∼9,000-20,000 cancer cells per subtype and ∼24,000 healthy cells for training, and ∼600-3,100 cells per subtype in the test sets. AML cell lines have been purchased from DSMZ. Cells were thawed in prewarmed medium (KASUMI1 – 80% RPMI 1640 + 20% h.i. FBS; MKPL1, MOLM13 and THP1 – 90% RPMI 1640 + 10% h.i. FBS; OCI-AML2 and OCI-AML3 – 90% alpha-MEM with ribo- and deoxyribonucleosides + 10% h.i. FBS; OCI-M2 - 80% Isocove’s MDM + 20% h.i. FBS) and immediately centrifugated at 400g for 5 min at 21°C. Cell lines were then cultured and allowed to recover for at least 2 days before performing imaging experiments. Cell lines were split every 2-3 days at 1:2 ratio. Peripheral blood mononuclear cells (PBMCs) from healthy donors were isolated from buffy coats obtained from the Zurich Blood Bank. Isolation was performed by density gradient centrifugation (SepMateTM-50, STEMCELL Technologies; Lympholyte-H, Cederlane). Aliquots of PBMCs were cryopreserved for subsequent use. After thawing, cells were cultured in RPMI 1640 and used directly for experiments. A total of 75 000 cells in 100 µL were plated per well in flat-bottom 96-well plates, either alone or at the indicated cell ratios. The plate was cultured under controlled conditions (37 °C, 5 % CO₂) in the stage top incubator during imaging using a Zaber automated brightfield microscope. Z-stack images were acquired at 20× magnification, with a fixed exposure time of 7 ms. Each stack consisted of 6 optical slices collected at 5 µm intervals and was centered on the cellular focal plane determined using the microscope’s autofocus feature, configured to maximize edge contrast while scanning in 10 µm increments over a 140 µm range. Each well was imaged every 6 hours for a total duration of 48 hours. At each time point, four non-overlapping fields of view were acquired per well in a grid pattern, with fields separated by 1000 µm in x and 1000 µm in y dimensions. All donors provided written informed consent, and all procedures were conducted in accordance with approval from the local ethics committee (BASEC 2020-00325).

Two patient-derived AML cohorts were also included in this work: the AML cohort 1 from the DECIDER^37^ study (25 patients, 203-434 cells per patient) and the AML cohort 2 (17 patients, 195-495 cells per patient) collected in the University Hospital Freiburg. Both cohorts were obtained from peripheral blood smears of patients at diagnosis, before initiation of therapy. EDTA-anticoagulated blood was Giemsa-stained and digitized using the CellaVision DC-1 system at a resolution of 360×360 pixels. The use was approved by the Ethics Committee of the Albert-Ludwigs-University Freiburg (Nr. 23-1567-S1-retro). All data were pseudonymized prior to analysis. The studies were conducted in accordance with the Declaration of Helsinki, and informed consent was obtained from all participants.

#### Open-source datasets

Label-free live-cell datasets included brightfield recordings of non-malignant breast epithelial cells^30^ (MCF10-A, n = 79,178). Stained open-source histopathology datasets comprised breast^31^ (BCSS-WSSS; 15,232 cancer, 8,190 healthy) and lung adenocarcinoma^31^ (LUAD; 13,151 cancer, 3,527 healthy) whole-slide images (H&E-stained). The BSCCM (Berkeley Single Cell Computational Microscopy) dataset^32^ provided LED-array brightfield images of healthy granulocytes, lymphocytes, and monocytes (n = 9,453). Stained white blood cell datasets included Raabin^33^ (n = 16,633), LISC^34^ (n = 257), Acevedo^35^ (n = 17,092), and Bodzas^36^ (n = 16,027), together covering a broad spectrum of leukocyte subtypes, both healthy and malignant, across distinct imaging conditions.

When datasets were not already available at the single-cell or patch level, individual cells were segmented using CellSAM^28^. Collectively, these datasets span label-free and stained imaging modalities, healthy and malignant phenotypes, and encompass hematologic and solid tumor contexts across multiple devices and institutions.

### *CellSign*: Framework for Generating CDFs

Ce*llSign* generates CDFs by transforming single-cell images into refined population-level representations. To do so, deep embeddings are computed using vision foundation models and adjusted with HCR. The resulting cell embeddings are then ordered along a continuous morphological trajectory and summarized with a dedicated pooling operator, yielding a fixed-length fingerprint for each population.

#### Deep Representation of Cell Populations using Vision Foundation Models

We propose a label-free, and hence unsupervised, pipeline that infers dynamic cell state structures (e.g., cell-cycle progression or myelopoiesis) from single-time-point microscopy images of cell populations. The approach consists of four main steps: segmentation, embedding, ordering, and pooling, resulting in a population dynamics fingerprint ℱ:

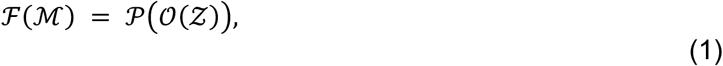

where

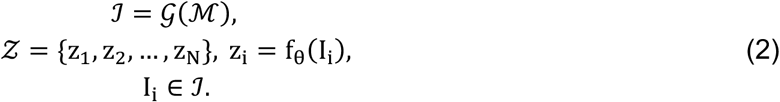

Here, ℳ = {M_1_,…, M_K_} denotes the set of raw microscopy images, 𝒢 is an instance segmentation operator that extracts single-cell images ℐ = {I_1_,…, I_N_}, *f*_*θ*_ is a parameterized function which maps each single-cell image to its latent embedding *z*_*i*_, 𝒪 orders these embeddings into trajectories, and 𝒫 pools the ordered embeddings into a fingerprint ℱ(ℳ) ∈ ℝ^h^. Thus, ℱ represents the full pipeline mapping from raw microscopy images through segmentation, embedding, ordering, and pooling. Finally, these fingerprints can be quantitatively compared using distance or similarity measures (e.g., Wasserstein distance) to reference fingerprints, enabling the identification of phenotypic shifts, disease-associated patterns, or treatment effects.

Each cell image I_i_ ∈ ℝ^H×W×C^ is passed through one or multiple (pretrained) deep neural network(s) to generate a non-linear latent representation z_i_ ∈ ℝ^d^ (also called embedding). Formally, we define

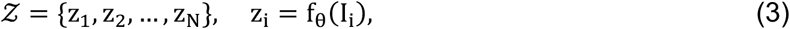

where *f*_*θ*_ is a parameterized function.

#### Healthy-Component Reduction (HCR)

Before ordering and pooling, we disentangle disease-related morphological structure from shared physiological variation by removing dominant sources of variance in cell morphology common to both healthy and malignant cells, such as the cell cycle, while preserving residual components that are indicative of pathological change. This procedure rests on the assumption that, at least in approximation, the cell-cycle pattern in malignant cells resembles that of healthy cells.

Given healthy cell embeddings X_healthy_ and malignant cell embeddings X_malignant_, we identify the principal subspace of healthy variation using PCA. Let U ∈ ℝ^d×k^ denote the top-*k* eigenvectors explaining a cumulative variance threshold τ (typically τ = 0.9). The corresponding projection matrix onto the healthy subspace is

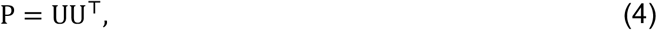

and the orthogonal complement, representing the reduced space, is

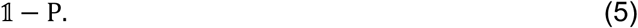

Each embedding x ∈ R^d^ is then transformed as

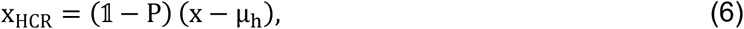

where µ_h_ is the mean of the healthy embeddings. This transformation retains only those morphological components that are orthogonal to the dominant healthy variation, thereby emphasizing differences associated with malignancy or dysregulation.

#### Cell Dynamics Fingerprints (CDFs)

After HCR, all embeddings are ordered into pseudotemporal trajectories and aggregated into CDFs. We decompose this process into two stages:

(i) Distance-based Sorting, which re-indexes cells into a latent trajectory (e.g., cell cycle progression),
(ii) Population Pooling, which compresses the ordered sequence into a fixed-length fingerprint.

#### Distance-based Sorting

We define an ordering operator 𝒪 to re-index the set of cell embeddings 𝒵 into a pseudotemporal sequence. Formally, we seek a permutation π ∈ 𝒮_𝒩_ such that:

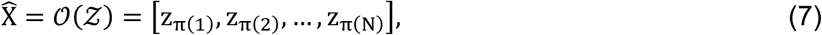

where the permutation π is determined according to a general criterion defined on a distance (or similarity) measure δ in embedding space:

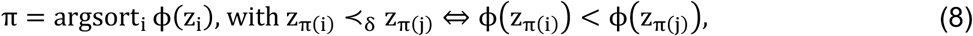

and **ϕ**: 𝒵 → ℝ is any scalarization derived from δ.

Here, we use PHATE which constructs a diffusion geometry over the high-dimensional embedding space and computes a low-dimensional embedding that preserves both local and global structure. It is particularly effective for data with branching or tree-like topologies, as the diffusion process naturally captures bifurcations and hierarchical relationships between states. We define a pseudotemporal ordering by projecting cell embeddings into the first diffusion component:

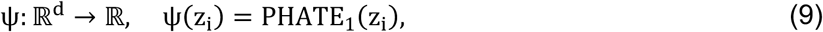

where PHATE_1_ denotes the first coordinate of the PHATE embedding, also called pseudotime. The ordered sequence is then obtained by sorting:

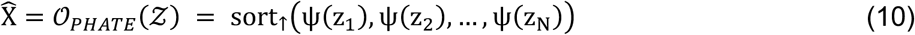

This approach captures smooth nonlinear trajectories by leveraging manifold structure, making it well-suited for gradual biological progressions such as differentiation or the cell cycle.

#### Population Pooling

Pooling then aggregates a list of ordered embeddings into a CDF *F* ∈ ℝ^h^ of a fixed length h. KDE pooling estimates the continuous density distribution of cell states along the ordered trajectory. Instead of taking simple statistical moments (e.g., mean or variance), KDE pooling models the full probability density of embeddings or their pseudotime coordinates, thereby capturing multimodal and asymmetric structures that reflect distinct subpopulations or state transitions. Formally, given the ordered embeddings 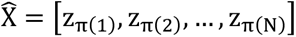, the pooled fingerprint is computed as

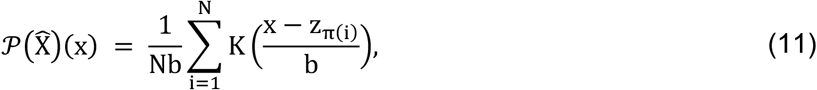

where K(⋅) is a kernel function (typically Gaussian), and b denotes the bandwidth parameter controlling smoothness. The resulting density 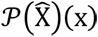 provides a continuous estimate of the embedding distribution and serves as a population-level representation that reflects how cells populate morphological state space over pseudotime. To obtain a compact fingerprint vector *F*, the density is evaluated on a fixed grid or projected onto a low-dimensional basis, yielding the population dynamics fingerprint.

### Performance Evaluation and Scoring

Performance was evaluated using macro-median ROC curves and corresponding macro-median AUROC values across repeated runs. For each run *r*, the ROC curve was computed by varying a discrimination threshold *t* over the malignancy scores *s*. At each threshold, the true-positive rate (TPR) and false-positive rate (FPR) were defined as:

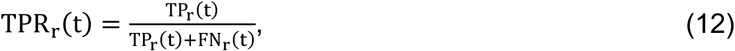

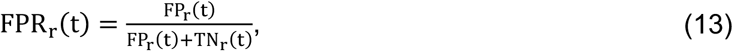

where TP_r_(t), FP_r_(t), TN_r_(t), and FN_r_(t) denote the counts of true positives, false positives, true negatives, and false negatives at threshold t in run *r*.

All ROC curves were interpolated on a shared grid of FPR values. The macro-median ROC curve was then defined as the pointwise median of the true-positive rates across runs:

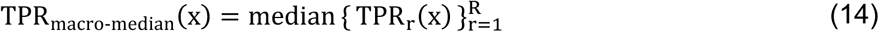

and the macro-median AUROC as the area under this curve:

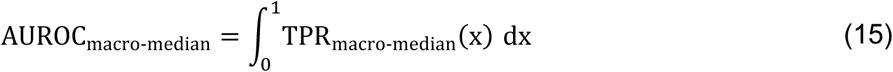

To quantify the malignancy of a given CDF *F*, we define a malignancy score based on its relative Wasserstein distances to two reference fingerprints: a healthy reference CDF *F*_healthy_ and a malignant reference CDF *F*_malignant_. Specifically, we compute

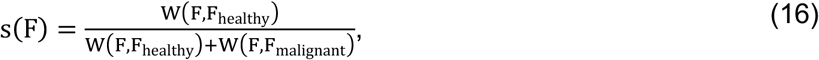

where *W*(⋅,⋅) denotes the Wasserstein distance between fingerprints. The score takes values in [0,1], yielding values closer to zero for CDFs that resemble the healthy reference and values closer to one for the CDFs that are closer to the malignant reference.

The Wasserstein distance *W*(*p*, *Q*) between one-dimensional distributions *p* and *Q* with cumulative distribution functions F_P_ and F_Q_ is given by:

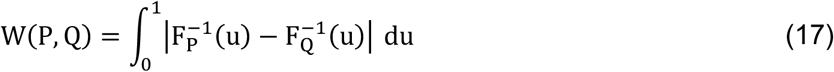

For plotting, scores were Z-normalized within each run:

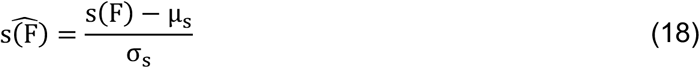

with µ_*s*_ and σ_*s*_ denoting the mean and standard deviation of all scores in that run.

### Hyperparameters

For ordering, dimensionality was first reduced to 32 principal components via PCA, followed by PHATE to obtain a one-dimensional manifold representation. The explained variance threshold for HCR was set between 80% and 99.9%, depending on dataset and transfer complexity. Kernel density estimates of the PHATE coordinates were computed on a fixed grid spanning −3 to 3 (55 points) using Gaussian kernels with dataset- and transfer-specific bandwidths: 1 (Kreutmair), 20 (breast histology and Bodzas+LISC-to-Kreutmair), 200 (lung histology), and 500 (AML Dataset 1 and BSCCM). Reference fingerprints for malignant and healthy populations were obtained as the mean of 30 runs, each using 200 randomly sampled cells. For transfer evaluations, 1,000 runs of 200 samples were used, while for patient-level datasets such as AML Dataset 1 and AML Dataset 2 all available cells per patient were included without subsampling.

## Notes

### Funding Statement

This research was funded by the Mertelsmann Foundation by the grant numbers Nr. 2025031 (to JDA) and MF-2024017 (to SK). SK was further supported by a fellowship for attending physicians from the Promedica Foundation.

### Author Declarations

Ethics Committee of the Canton of Zurich gave ethical approval for this work involving the isolation and imaging of peripheral blood mononuclear cells from healthy donors (BASEC 2020-00325). All donors provided written informed consent. Ethics Committee of the Albert-Ludwigs-University Freiburg gave ethical approval for this work involving the retrospective analysis of peripheral blood smears from patients with acute myeloid leukemia (protocol number 23-1567-S1-retro). All data were pseudonymized prior to analysis, and informed consent was obtained from all participants.

